# Effect of uni- and multi-modal exercise interventions on visuo-motor functional connectivity in glaucoma patients and healthy elderly

**DOI:** 10.64898/2025.12.17.25342381

**Authors:** Rohit Misra, Gokulraj T Prabhakaran, Mahima V Rebello, Khaldoon O Al-Nosairy, Rosalie Beyer, Constantin Freitag, Cynthia Moffack Djuloun, Francie H Stolle, Martin Behrens, Tom Behrendt, Hagen Thieme, Lutz Schega, Michael B Hoffmann

## Abstract

**Motivation:** Glaucoma is a progressive visual pathway disorder leading to visual field loss, which can be linked to disruptions in visuo-motor function and an increase in fall risk. Motor-cognitive training has been shown to affect visuo-motor functional connectivity (FC) in healthy controls (HC) and could therefore be a viable strategy for the management of glaucoma patients (GL).

**Methods:** Using 3T resting state functional-MRI (rs-fMRI), we studied the effect of a multimodal intervention (MMI [resistance + motor-cognitive dual-task training]) and unimodal intervention (UMI [resistance training only]) on rs-FC in 20 HC and 12 GL, randomly assigned to MMI (7GL, 9HC) and UMI (5GL, 11HC). Rs-fMRI was acquired pre- and post the 12-week intervention. Whole-brain FC were estimated with bilateral seeds in the visuo-motor pathway and paired longitudinal comparisons were performed to test effects of the interventions.

**Results:** Longitudinal effects of UMI were only observed in HC and were limited to an increase in FC between the cerebellum, midcingulate gyrus, and the frontal eye fields (pFWE<0.05). Interestingly, MMI had a widespread effect including an increase in FC within the visual pathway, between auditory and visual cortices, and with the motor cortex after MMI (pFWE<0.05) in both HC and GL. In addition, an increased FC between the dorsolateral prefrontal cortex, inferior frontal gyrus, sensorimotor cortex, and the inferior temporal gyrus (pFWE<0.05) was observed in GL after MMI. An increase in FC was also found between the cerebellum and the midcingulate gyrus (pFWE<0.05) in GL that underwent MMI.

**Conclusion:** Overall, our findings indicate a widespread effect of MMI in both HC and GL on rs-FC, highlighting cortical plasticity in GL. These results motivate further research on the effects of motor-cognitive interventions for visual pathway disorders like glaucoma.

## 1 Introduction

Visuo-motor function is essential for everyday activities such as walking and relies on continuous interaction between sensory, motor, and cognitive systems. Aging naturally affects these systems, e.g., by reducing muscle strength (Izquierdo et al., 2021; Clark & Manini, 2012; Mau-Moeller et al., 2013; Nguyen et al., 2023) and impairing executive functions (Cohen et al., 2016), which together contribute to gait instability and an increased fall risk (MacAulay et al., 2015; Fragala et al., 2019). Importantly, falls are the leading cause of injury in the elderly population (Statistisches Bundesamt, 2023–24). If the age-related impairments are accompanied by diseases that disrupt sensory input, particularly vision, the potential for gait impairment and falls increases (Freitag et al., 2023). Therefore, the development of interventions that can support or restore visuo-motor functioning in older adults with vision disorders is of particular importance.

Glaucoma is a visual pathway disorder that presents with progressively worsening visual field defects and is the leading cause of irreversible blindness worldwide (Steinmetz et al., 2021). The disease involves chronic progressive neuropathy of the optic nerve, rooted in the loss of retinal ganglion cells in the retina and optic nerve (Jonas et al., 2000). Glaucomatous damage, however, is not limited to the eyes but also shown to affect brain structure and function (reviewed in Nuzzi et al., 2018, 2021). In addition to the risks associated with healthy aging, glaucoma presents with impaired peripheral vision and orientation difficulties (Andac et al. 2024), which further increase the fall risk in these patients (Jian-Yu et al., 2021). It has also been observed that reduced balance performance in glaucoma patients is correlated with impairment in sensorimotor brain connectivity (Trivedi et al., 2019). This suggests that gait dysfunction in glaucoma may not only be linked to the eyes but is also associated with aberrations in the visuo-motor coordination circuits in the cortex.

Targeting neuromuscular function with resistance training may counteract age-related declines in sensory-motor function (Fragala et al., 2019) and can indirectly support cognitive improvements (Herold et al., 2019; Liu-Ambrose et al., 2008). Building on this, interventions that combine motor and cognitive training have shown promise in mitigating gait and balance impairments in elderly populations (Wollesen et al., 2014, Schoene et al., 2014) and in neurological disorders such as Parkinson’s disease and mild cognitive impairment (Allen et al., 2011, Dove et al., 2020). The Life Kinetik® program is one such approach that combines physical training with cognitively demanding tasks that challenge sensory integration, working memory, attention, and executive functions (Lutz 2021). In healthy adults, this multimodal intervention has been demonstrated to induce widespread changes in resting-state functional connectivity (FC), including increased coupling between visual, auditory, and motor cortices (Demirakca et al., 2016). However, whether such approaches can address visuo-motor deficits in glaucoma remains unknown.

From a neuroimaging perspective, studies using resting-state functional magnetic resonance imaging (fMRI) in glaucoma have observed disruptions in functional connectivity between visual cortices and regions involved in visuo-motor control, such as the motor cortex, cerebellum, and parietal association areas, potentially contributing to deficits in gait stability and spatial navigation (Trivedi et al., 2019). Resting-state FC might, therefore, also be of value as a non-invasive marker for detecting cortical reorganization induced by various intervention schemes. By assessing resting-state FC before and after targeted interventions, it may be possible to capture plasticity within sensorimotor networks that may underlie functional improvements (Demirakca et al., 2016; Nuzzi et al., 2021). To our knowledge, no prior study has evaluated the impact of motor–cognitive training or compared it to unimodal motor training on FC in glaucoma.

In this study, we examined the effect of a multimodal intervention (MMI [motor-cognitive dual-task training + resistance training]) and unimodal intervention (UMI [resistance training only]) on resting-state FC in glaucoma patients (GL) and healthy controls (HC). Based on findings by Demirakca et al. (2016) in HC, we used a seed-based approach to examine longitudinal changes in whole-brain FC of regions implicated in visual processing, auditory processing, and motor coordination. We hypothesized that (i) MMI would induce more widespread changes in FC compared to UMI in both HC and GL, (ii) MMI would induce FC changes in GL in regions involved in multisensory integration as a compensatory mechanism for reduced visual input, and (iii) with UMI, we expected to see FC changes primarily in regions involved in motor planning and coordination.

## 2 Methods

### 2.1 Study Design

In this two-arm, randomized controlled study, we aimed to investigate the effect of two different intervention schemes on resting-state FC in patients with early-to-moderate stage GL and a reference group of HC. The HC and GL were randomly assigned to two groups, respectively, that underwent MMI and UMI for 12 weeks. Clinical ophthalmological measurements as well as brain MR imaging were conducted before and after the interventions (German Clinical Trial Register, ID: DRKS00022519/05.08.2020, https://drks.de/search/de/trial/DRKS00022519). Study procedures were approved by the Ethics Committee of the University Medical Faculty Magdeburg (32/18) and conducted in accordance with the principles outlined in the Declaration of Helsinki. Reporting was performed in accordance with the Consolidated Standards of Reporting Trials (CONSORT) Statement for randomized pilot trials (Eldridge et al., 2016). All participants signed the informed consent form before participating in this study.

### 2.2 Participants and Recruitment

Participants were recruited at the Department of Ophthalmology at the University Hospital Magdeburg through local ophthalmologists and the national patient network. The sample size was oriented towards the study by Demirakca et al. (2016), who reported significant effects of the motor-cognitive Life Kinetik® intervention (21 healthy adults; intervention participants) compared to a control group (11 healthy adults; no intervention) on FC. It should be noted that data collection and funding for our study took place during the COVID-19 pandemic, which limited recruitment and resulted in a total sample of 32 participants.

The criteria to be included in the study were: (i) age ≥ 60 years, (ii) diagnosis of open-angle glaucoma (for the GL cohort), and (iii) ability to walk at least 6 minutes without walking support. The exclusion criteria were: (i) diagnosis of any eye disease affecting measures of visual function (e.g. cataract [except incipient stage], history of ocular trauma or ocular surgeries other than glaucoma of cataract surgery), (ii) neurological diseases, and (iii) diseases that limit the physical performance of the participants (orthopedic diseases like arthrosis [grade II or above], musculoskeletal impairments, tendinitis, tenosynovitis, myositis, prosthesis in lower extremities, joint replacements, neurological disorders, rheumatism, cardiovascular disorders, stroke). Further, participants who did not attend more than 80% of the sessions (i.e., less than 19.2 hours) were excluded from this study. Allocation was carried out at the Department of Sport Science. Participants were randomly assigned to either the MMI or UMI group using counterbalanced randomization (allocation ratio 1:1) based on a computer-generated random number table. After visual inspection of the MRI data and checking for acquisition artifacts, a total of 20 HC (72.2 ± 5.5 years, 10 females) and 12 GL (70.2 ± 4.6 years, 5 females) were included in this study. A flowchart detailing the recruitment procedure is shown in Figure 1.

**Figure 1.**
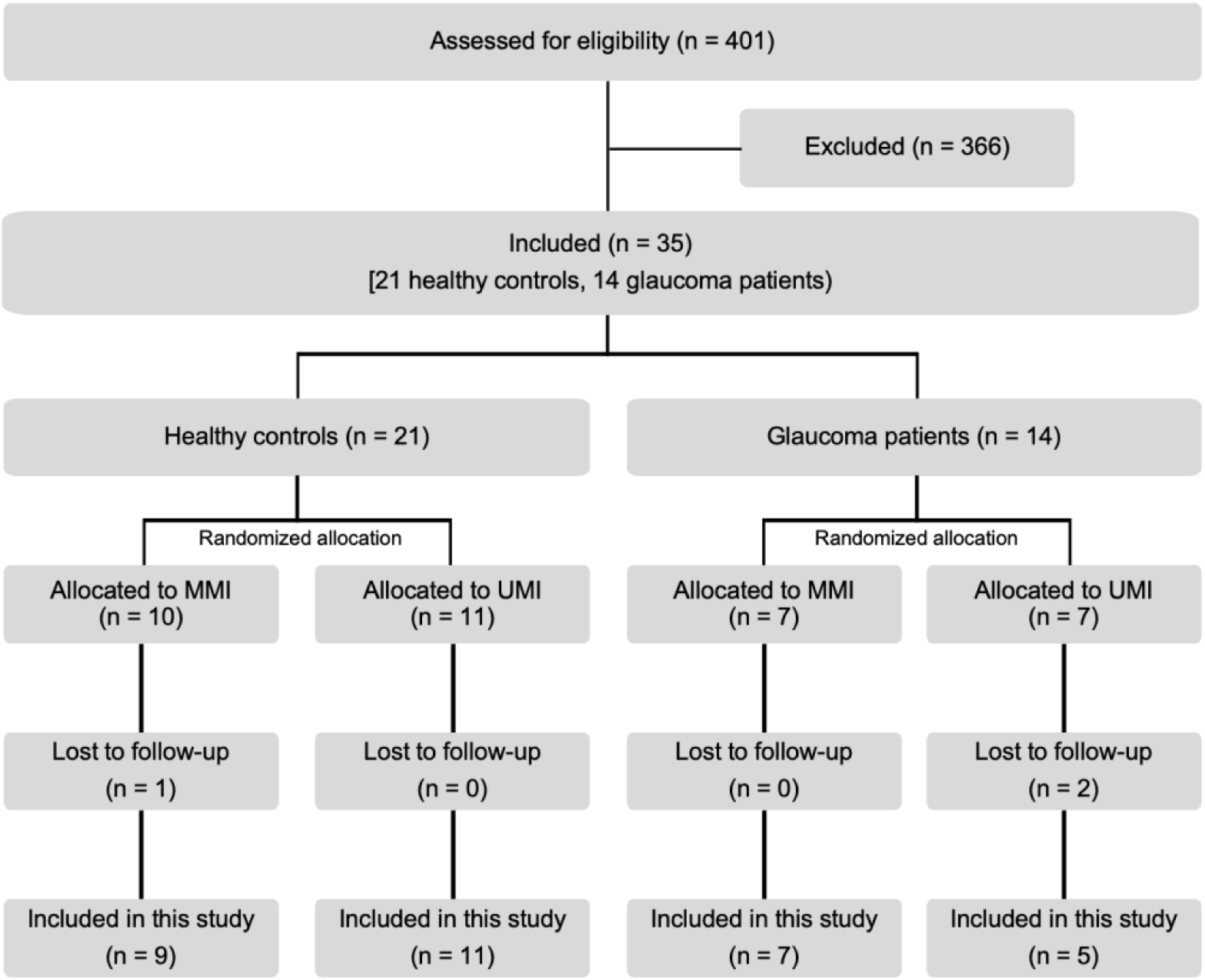
Flowchart of the recruitment procedure for the study. MMI = multimodal intervention, UMI = unimodal intervention.

#### 2.2.1 Clinical Assessment

Ophthalmic examinations of all participants were conducted at the ophthalmology department at the University Hospital, Magdeburg from September 2020 - December 2022. The participants underwent the following tests: Best-corrected visual acuity (BCVA) testing, stereovision testing with the Lang test, visual field (VF) testing (24-2 SITA-Fast test, Humphrey Field Analyzer, Carl Zeiss Meditec AG, Jena, Germany), peripapillary retinal nerve fiber layer thickness with optical coherence tomography (OCT, Heidelberg Engineering, Heidelberg, Germany), and retinal fundus examination (details published in Beyer et al., 2024). Ocusweep was used for binocular VF testing in an ambient room light at ∼40 cm distance (Ocusweep^®^, Ocuspecto Ltd, Turku, Finland).

#### 2.2.2 Interventions

The interventions were performed in the laboratories of the Sport Science Department at the Otto von Guericke University Magdeburg. The participants were divided into two groups to undergo different intervention schemes, with 16 participants in each group: (a) MMI (7 GL, 9 HC; 6 females, 10 males; 72.3 ± 5.7 years) and (b) UMI (5 GL, 11 HC; 9 females, 7 males; 70.7 ± 4.7 years). Table 1 summarizes the demographic details of the sub-groups for each intervention. Both intervention groups participated in a 12-week training program consisting of two sessions per week (24 sessions in total) conducted on non-consecutive days. Each session lasted 60 minutes and was supervised by experienced instructors.

**Table 1.** Demographic characteristics of the groups.

|  | Healthy controls (HC) |  |  | Glaucoma patients (GL) |  |  |
| --- | --- | --- | --- | --- | --- | --- |
|  | All HC | HC - UMI | HC - MMI | All GL | GL - UMI | GL - MMI |
| Number of participants | 20 | 11 | 9 | 12 | 5 | 7 |
| Age (mean ± std, in years) | $72.2 \pm 5.5$ | $71.0 \pm 5.4$ | $73.8 \pm 5.6$ | $70.2 \pm 4.6$ | $70.0 \pm 3.2$ | $70.4 \pm 5.6$ |
| Sex (M/F) | 10/10 | 6/5 | 4/5 | 7/5 | 1/4 | 6/1 |

*M = male, F = female, HC = healthy controls, GL = glaucoma patients, UMI = unimodal intervention, MMI = multimodal intervention, std = standard deviation*

Before the interventions started, details regarding daily physical activity, age, height, and weight were collected. Further, the pre-intervention assessment included measurements of visual, cognitive, and motor function as well as spatio-temporal gait parameters (Lutz 2021, Freitag et al. 2023, Freitag et al. 2024). Disease progression was assumed to be negligible during the intervention period. Both interventions are described in detail by Freitag et al. (2025). Shortly, the MMI intervention included motor-cognitive dual-task training based on the Life Kinetik® program combined with resistance training, whereas the UMI group performed resistance training only. Resistance exercises were paced using a metronome set at 60 bpm (7.5 repetitions; 2 seconds eccentric / 2 seconds concentric; 2 sets per exercise) and were performed with free weights and exercise machines in a fixed sequence. To avoid increases in intraocular pressure associated with supine positions, all exercises were carried out in a seated position as exercising in a supine position has been shown to affect the intraocular pressure (Najmanová et al. 2010, Lara et al. 2023).

### 2.3 MRI Acquisition

The MRI measurements were performed with a 3T Siemens Prisma scanner (Erlangen, Germany) at the University Hospital Magdeburg. The structural T1-weighted images were acquired using the MPRAGE protocol (1 mm isotropic voxels, TR | TI | TE = 2500 | 1100 | 2.82 ms). The resting-state functional MRI (rs-fMRI) scans were acquired using a T2*-weighted BOLD gradient-EPI sequence (TR | TE = 1500 | 30 ms & voxel size = 2.5^3^ mm^3^). Two runs of rs-fMRI were acquired, each lasting 6 minutes (i.e. 240 measurements/run × 2 runs). During the run, the participants were instructed to lie as still as possible and, in accordance with Demirakca et al. (2016), with their eyes open and to not fall asleep. A blank black screen with a white fixation cross was presented during the scan. The fixation of their dominant eye (in GL: based on VF testing, in HC: using Miles test) was tracked to monitor if they were falling asleep. In case the participant fell asleep, the acquisition was stopped, and the run was started again.

### 2.4 Data Preprocessing

The data were processed in MATLAB (Mathworks, R2022a) using the statistical parametric mapping (SPM12) (The Wellcome Centre for Human Neuroimaging, 2023, http://www.fil.ion.ucl.ac.uk/spm/software/spm12/) and CONN toolboxes (Whitfield-Gabrieli & Nieto-Castanon, 2012; Nieto-Castanon & Whitfield-Gabrieli, 2021; Penny et al., 2011). The anatomical images were segmented into grey matter, white matter, and cerebro-spinal fluid (CSF) using SPM12. Preprocessing of the functional scans involved motion correction, co-registration, DARTEL-based normalization, smoothing, denoising, and temporal filtering. Motion correction was performed by realigning volumes of both runs of the participant to the first volume of the first run. None of the scans had framewise displacement of more than 2 millimeters due to motion. So, no scans were rejected due to excessive motion. The fMRI volumes were co-registered to their anatomical images using a 6-parameter rigid body transformation. The segmented gray and white matter images of all participants in the study were used to create a study-specific template using DARTEL (Ashburner, 2007), which was then normalized to MNI space; both anatomical and functional images were normalized to this template. The functional scans were smoothed using a Gaussian kernel with full-width half maximum (FWHM) of 8 mm. Denoising was performed using the anatomical CompCor approach (Behzadi et al., 2007; Chai et al., 2012). The first 5 principal components each of the white matter and CSF, along with the 6 motion regressors, were regressed out of the BOLD time series. Finally, a bandpass filter with cut-offs at 0.01 Hz and 0.1 Hz was used for temporal filtering. As the fMRI slices were acquired in an interleaved manner and the TR was small, we did not perform slice time correction for the functional data.

### 2.5 Seed-Based Functional Connectivity

For performing a seed-to-voxel FC analysis, we selected the seed regions based on the targeted brain functions discussed earlier. Based on the results of Demirakca et al. (2016) on the effect of Life Kinetik® on FC in healthy controls, regions of the brain expected to be involved in functions such as primary sensory processing, visuo-motor coordination, motor planning, multisensory integration, and executive functions were selected as seeds. We selected the corresponding Brodmann Areas (BA) using the Wake Forest University (WFU) PickAtlas (Maldjian et al., 2003, 2004) (http://fmri.wfubmc.edu/software/PickAtlas). The seeds included bilateral masks of the BA1 (primary sensorimotor cortex), BA4 (primary motor cortex), BA9 (dorsolateral prefrontal cortex), BA17 (primary visual cortex), BA18 (secondary visual cortex), BA41 (primary auditory cortex), and BA42 (secondary auditory cortex). Additionally, masks for the right and left frontal eye fields (FEF) and Mid-Cingulate Gyrus (MCG) were created using the HCP Multi-Modal Parcellations atlas (Glasser et al., 2016). Overall, a total of 18 seeds were studied (Figure 2).

**Figure 2.**
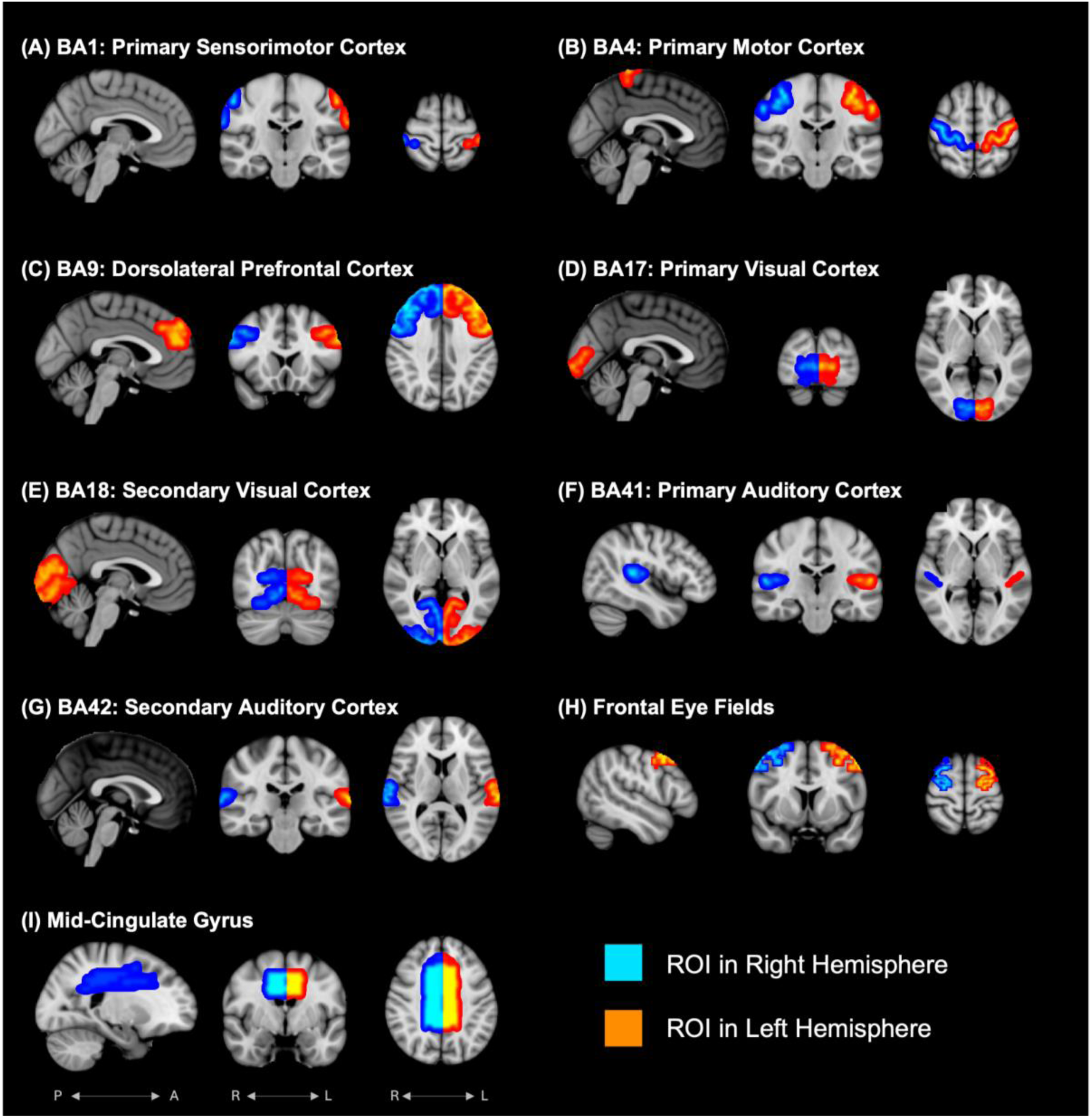
Regions of Interest (ROIs) used as seeds for seed-based functional connectivity analysis. (A) Primary sensorimotor cortex (BA1), (B) Primary motor cortex (BA4), (C) Dorsolateral prefrontal cortex (BA9), (D) Primary visual cortex (BA17), (E) Secondary visual cortex (BA18), (F) Primary auditory cortex (BA41), (G) Secondary auditory cortex (BA42), (H) Frontal eye fields (FEF), and (I) Mid-cingulate gyrus (MCG). BAxx = Brodmann Area, P = Posterior, A = Anterior, R = Right, L = Left..

For each seed mask, the representative time series was obtained by averaging the time series of all the component voxels in that seed. Seed-based FC maps were generated by calculating the Pearson’s correlation coefficient between the representative time series and the time series from all other voxels in the brain. The correlation values were Fisher-*z* transformed to facilitate normality for further statistical analyses.

### 2.6 Statistical Analysis

First, we checked for baseline differences between the HC and GL groups irrespective of the intervention method. To test this, seed-based FC maps for all GL pre-intervention were compared with HC pre-intervention using a two-sided two-sample t-test. Age and sex of the participants were added as covariates of no interest in the model. To obtain significant clusters in the tests, a cluster-forming threshold of p < 0.001 (uncorrected) was used, followed by correction for multiple comparisons using a Family-Wise Error (FWE) of p_FWE_ < 0.05.

A complete 2^3^ factorial model (with Group (HC/GL), Intervention (UMI/MMI), and Time (Pre/Post) as factors) was not applied due to the small sample sizes in the 4 groups (i.e., MMI and UMI for both HC and GL, respectively). The high inter-subject variability in FC would make unpaired comparisons uninterpretable. So, we limited the scope of the statistical analyses to pairwise longitudinal comparisons within each group. Therefore, here we focus primarily on the longitudinal effects of the intervention methods on the respective groups. For each of the four sub-groups, (i) HC that underwent UMI (ii) HC that underwent MMI, (iii) GL that underwent UMI, and (iv) GL that underwent MMI, we conducted paired two-sample t-tests to compare the FC maps pre- and post-intervention. Age and sex were added as covariates of no interest. To obtain significant clusters in the tests, a cluster-forming threshold of p < 0.001 (uncorrected) was used, followed by correction for multiple comparisons using a Family-Wise Error (FWE) of p_FWE_ < 0.05.

## 3 Results

### 3.1 Demographics and Ophthalmological Characteristics

After visual inspection of the MRI data and checking for acquisition artifacts, a total of 20 HC (72.2 ± 5.5 years, 10 females, 10 males) and 12 GL (70.2 ± 4.6 years, 5 females, 7 males) were included in this study. Table 1 provides demographic details of the cohorts and the sub-cohorts assigned for MMI or UMI. An unpaired two-sample t-test comparing the age distributions of the cohorts shows that the cohorts are age-matched (*p* = 0.3008). Chi-square test on the sex distributions of HC and GL cohorts was not significant with ***χ***^2^ = 0.2092 and *p* = 0.647 (*p* > 0.05). The patient ophthalmological characteristics are presented in Table 2.

**Table 2.** Ophthalmological characteristics of the cohorts.

|  | Healthy controls (N = 20) | Glaucoma patients (N = 12) |
| --- | --- | --- |
| MD of right eye in dB (median \| range) | -0.13 \| 5.09 | -0.36 \| 14.02 |
| MD of left eye in dB (median \| range) | 0.00 \| 5.92 | -1.04 \| 22.0 |
| **MD binocular in dB (median \| range)** | -0.7 \| 3.3 | 0.5 \| 9.6 |
| **BCVA in logMAR (mean ± std)** | -0.09 ± 0.08 | -0.08 ± 0.15 |
| **pRNFL right eye in $\mu\text{m}$ (mean ± std)** | 93 ± 12.78 | 79 ± 14 |
| **pRNFL left eye in $\mu\text{m}$ (mean ± std)** | 91 ± 11.19 | 76 ± 15 |

*BCVA = best corrected visual acuity [logMAR], pRNFL= peripapillary retinal nerve fiber layer (pRNFL) thickness [ $\mu\text{m}$ ], MD = mean deviation, std = standard deviation, decibel [db] (negative values indicate sensitivity reduction).*

### 3.2 Pre-Training Differences

Comparison of the FC maps of HC and GL before training, irrespective of the intervention scheme, showed significant differences (p_FWE_ < 0.05) for seeds in the left auditory association cortex (BA42), and the right frontal eye field (FEF) (see Figure 3 and Table 3). With the seed in auditory association cortex, significant clusters were observed in the right and left lateral occipital cortex (p_FWE_ < 0.05, GL HC, cluster size = 318 voxels and p_FWE_ < 0.05, GL > HC, cluster size = 210 voxels, respectively), in the right superior visual cortex (p_FWE_ < 0.05, GL > HC, cluster size = 159 voxels), and the medial orbitofrontal cortex (p_FWE_ < 0.05, GL > HC, cluster size = 213 voxels). With the seed in the FEF, we observed significant differences in its FC with the left precentral gyrus (p_FWE_ < 0.05, GL < HC, cluster size: 171 voxels).

**Figure 3.**
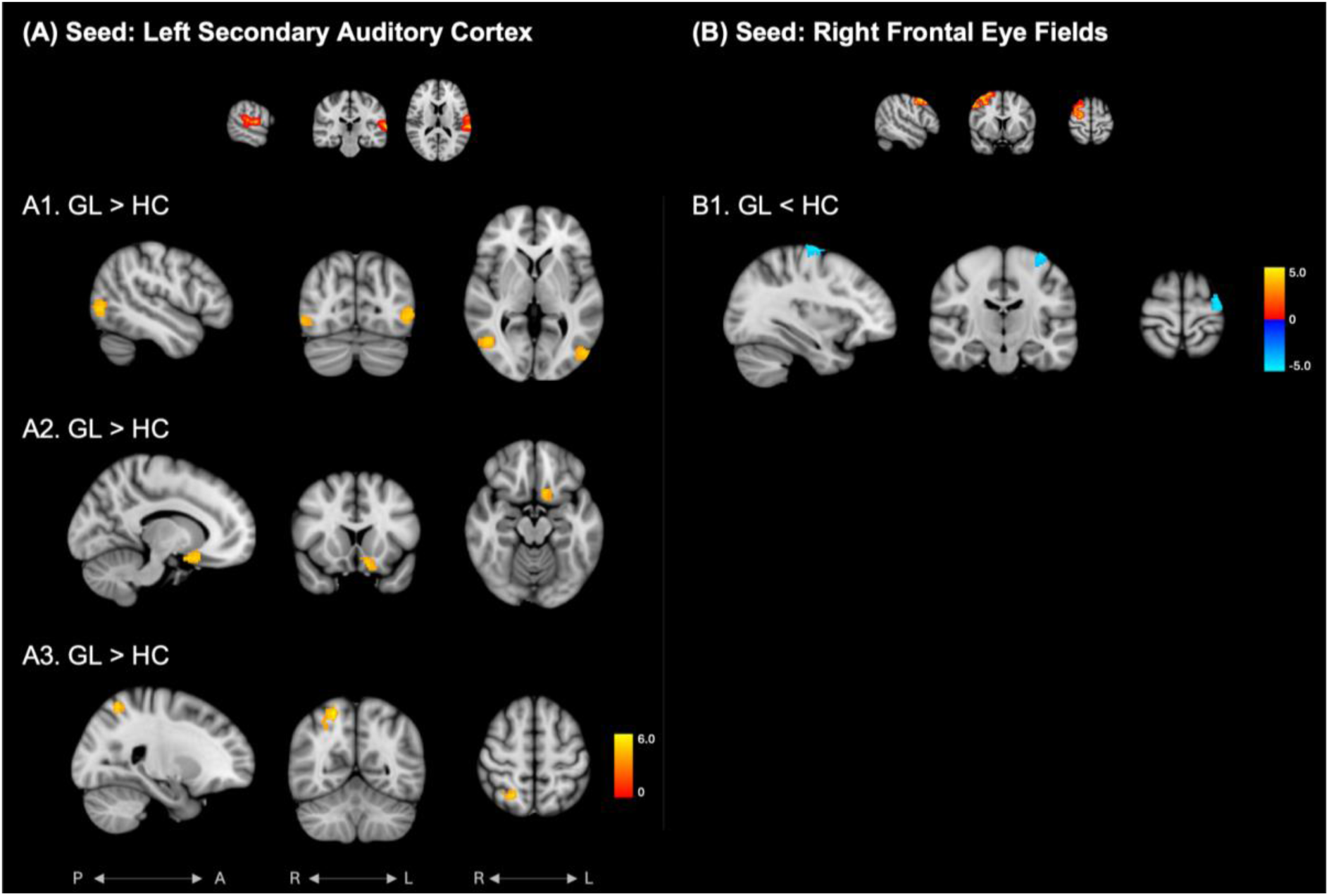
Significant differences between glaucoma (GL) and healthy controls (HC) before the interventions (pFWE < 0.05). (A) With seed in the left secondary auditory cortex, 3 clusters were observed at (A1) right and left lateral occipital cortex (GL HC), (A2) right superior visual cortex (GL > HC), and (A3) medial orbitofrontal cortex (GL > HC). (B) With seed in the right frontal eye fields, one cluster was observed in the (B1) left precentral gyrus (GL < HC). For details see Table 3. Colorbars represent t-values. FWE = Family-Wise Error rate.

**Table 3.** Summary of significant clusters (pFWE < 0.05). (A) Pre-training differences between healthy controls and glaucoma. (B) Effect of unimodal intervention in healthy controls. (C) Effect of multi-modal intervention in healthy controls. (D) Effect of multi-modal intervention in glaucoma. DOF = Degrees of Freedom, FWE = Family-Wise Error rate, “Min. pFWE” refers to the minimum corrected p-value within the cluster.

| (A) Pre-training differences between healthy controls and glaucoma [N = 32, DOF = 29] |  |  |  |  |  |  |  |
| --- | --- | --- | --- | --- | --- | --- | --- |
| Seed region | Cluster region | Contrast | Peak X (MNI, cm) | Peak Y (MNI, cm) | Peak Z (MNI, cm) | Cluster size (voxels) | Min. $p_{FWE}$ |
| Left secondary auditory cortex (BA42) | Right lateral occipital cortex | GL > HC | 46 | -66 | 4 | 318 | 0.0014 |
|  | Left lateral occipital cortex | GL > HC | -48 | -76 | 0 | 210 | 0.0137 |
|  | Right superior visual cortex | GL > HC | 22 | -58 | 58 | 159 | 0.0458 |
|  | Medial orbitofrontal cortex | GL > HC | -12 | 8 | -18 | 213 | 0.0128 |
| Right frontal eye fields | Left precentral gyrus | HC > GL | -38 | -18 | 70 | 171 | 0.0361 |
| (B) Effect of unimodal intervention in healthy controls (pre-training vs post-training) [N = 11, DOF = 8] |  |  |  |  |  |  |  |
| Seed region | Cluster region | Contrast | Peak X (MNI, cm) | Peak Y (MNI, cm) | Peak Z (MNI, cm) | Cluster size (voxels) | Min. $p_{FWE}$ |
| Left mid-cingulate gyrus | Right cerebellum | Post > Pre | 20 | -50 | -58 | 107 | 0.0074 |
| Left frontal eye fields | Right cerebellum | Post > Pre | 12 | -62 | -56 | 99 | 0.0237 |
| (C) Effect of multi-modal intervention in healthy controls (pre-training vs post-training) [N = 9, DOF = 6] |  |  |  |  |  |  |  |
| Seed region | Cluster region | Contrast | Peak X (MNI, cm) | Peak Y (MNI, cm) | Peak Z (MNI, cm) | Cluster size (voxels) | Min. $p_{FWE}$ |
| Left frontal eye fields | Right temporal occipital junction | Pre > Post | 64 | -58 | 0 | 68 | 0.0190 |
| Left primary motor cortex | Left insular cortex | Post > Pre | -38 | 18 | -12 | 82 | 0.0076 |
| Left primary visual cortex | Left visual association cortex | Post > Pre | -14 | -90 | 12 | 108 | 0.0016 |
| Left primary auditory cortex | Left parieto-occipital cortex | Post > Pre | -40 | -86 | 32 | 85 | 0.0036 |
|  | Left hippocampus, left parahippocampal gyrus | Post > Pre | -20 | -32 | -20 | 77 | 0.0067 |
| Left secondary auditory cortex | Left lateral occipital cortex | Post > Pre | -42 | -86 | 0 | 79 | 0.0065 |
|  | Right lateral occipital cortex | Post > Pre | 42 | -66 | -8 | 69 | 0.0145 |
| (D) Effect of multi-modal intervention in glaucoma (pre-training vs post-training) [N = 7, DOF = 4] |  |  |  |  |  |  |  |
| Seed region | Cluster region | Contrast | Peak X (MNI, cm) | Peak Y (MNI, cm) | Peak Z (MNI, cm) | Cluster size (voxels) | Min. $p_{FWE}$ |
| Left sensorimotor cortex | Left inferior frontal gyrus | Post > Pre | -52 | 30 | -2 | 52 | 0.0010 |
| Right primary motor cortex | Medial cuneal cortex / precuneus | Post > Pre | 2 | -86 | 32 | 41 | 0.0059 |
| Right dorsolateral prefrontal cortex | Left superior frontal gyrus | Post > Pre | -8 | 38 | 44 | 34 | 0.0074 |
|  | Left inferior frontal gyrus | Post > Pre | -52 | 28 | 18 | 29 | 0.0202 |
|  | Right inferior temporal gyrus | Pre > Post | 48 | -31 | -16 | 26 | 0.0379 |
| Right primary visual cortex | Left inferior temporal gyrus | Post > Pre | -64 | -46 | -18 | 30 | 0.0300 |
| Left secondary visual cortex | Right postcentral gyrus | Post > Pre | 28 | -32 | 64 | 69 | <0.0001 |
| Right primary auditory cortex | Left cuneal cortex | Post > Pre | -4 | -86 | 32 | 37 | 0.0063 |
| Left secondary auditory cortex | Left parieto-occipital cortex | Post > Pre | -14 | -76 | 52 | 41 | 0.0048 |
| Left mid-cingulate gyrus | Right cerebellum | Post > Pre | 48 | -70 | -36 | 32 | 0.0097 |
|  | Right precentral gyrus | Post > Pre | 58 | -6 | 20 | 31 | 0.0119 |
|  | Right cerebellum | Post > Pre | 26 | -80 | 28 | 29 | 0.0180 |

### 3.3 Effect of the Unimodal Intervention in Healthy Controls and Glaucoma Patients

Pairwise longitudinal comparisons within the group of GL that underwent UMI did not reveal any clusters with significant changes in the seeds tested. In contrast, within the group of HC that underwent UMI, we observed significant increase in the FC of two seeds, i.e., left MCG and left FEF, both with the right cerebellum (see Figure 4 and Table 3B).

**Figure 4.**
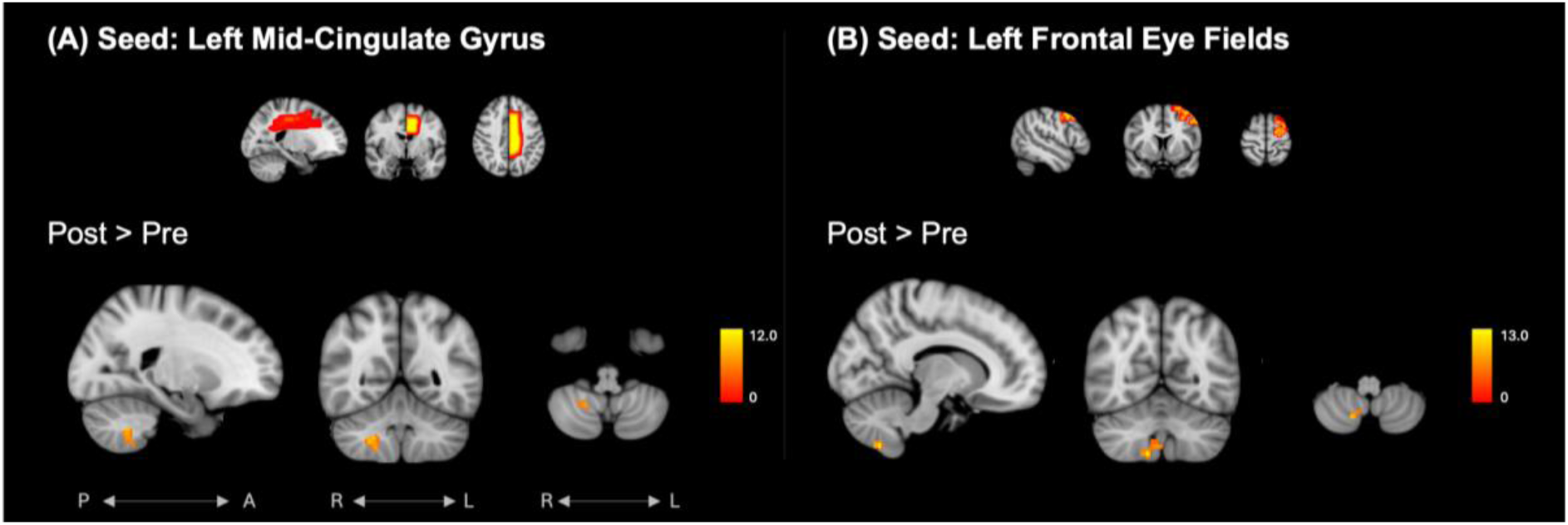
Significant differences in healthy controls before (Pre) and after (Post) the unimodal intervention (pFWE <0 .05). (A) With seed in the midcingulate gyrus, one cluster was observed at the right cerebellum (Post > Pre). (B) With seed in the left frontal eye fields, one cluster was observed in the right cerebellum (Post > Pre). For details see Table 3. Colorbars represent t-values. FWE = Family-Wise Error rate

### 3.4 Effect of the Multi-Modal Intervention in Healthy Controls

We tested the changes in rs-FC in HC that underwent MMI with longitudinal comparisons using paired t-tests. rs-FC changes were observed for five seeds, i.e., left primary and secondary auditory cortex, left frontal eye fields, left primary motor cortex, and left primary visual cortex (see Figure 5 and Table 3C). The left primary auditory cortex (BA41) revealed an increase in FC with the left parieto-occipital cortex (p_FWE_ < 0.05, Post > Pre, cluster size = 85 voxels, p_FWE_ = 0.0036) and the left hippocampus / parahippocampal gyrus (p_FWE_ < 0.05, Post > Pre, cluster size = 77 voxels, p_FWE_ = 0.0067). The FC between the left secondary auditory cortex (BA42) and the left (p_FWE_ < 0.05, Post > Pre, cluster size = 79 voxels, p_FWE_ = 0.0065) and the right lateral occipital cortices (p_FWE_ < 0.05, Post > Pre, cluster size = 69 voxels, p_FWE_ = 0.0145) showed an increase after MMI. The left FEF showed a significant decrease in FC with the right temporal occipital junction (close to the middle temporal gyrus) (p_FWE_ < 0.05, Post < Pre, cluster size = 68 voxels, p_FWE_ = 0.0190). The left primary motor cortex (BA4) showed altered FC with the left insular cortex upon training (p_FWE_ < 0.05, Post > Pre, cluster size = 82 voxels, p_FWE_ = 0.0076). FC between the left primary visual cortex (seed) and the left visual association cortex was also increased after MMI in HC (p_FWE_ < 0.05, Post > Pre, cluster size = 108, p_FWE_ = 0.0016).

**Figure 5.**
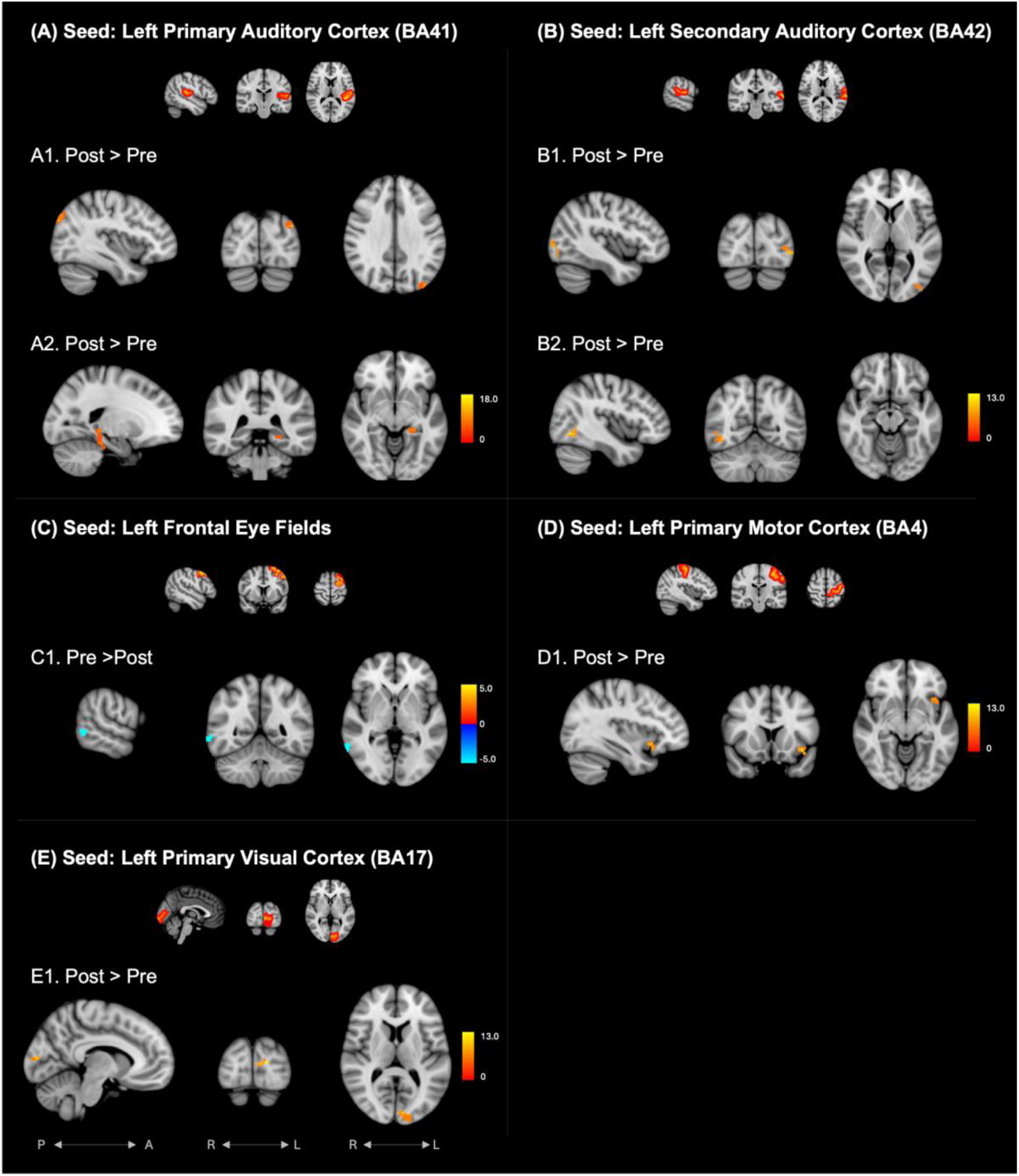
Significant differences in healthy controls before (Pre) and after (Post) the multi-modal intervention (pFWE < 0.05). (A) With seed in the left primary auditory cortex, two clusters were observed at (A1) left parieto-occipital cortex (Post > Pre) and (A2) left hippocampus / parahippocampal gyrus (Post > Pre). (B) With seed in the left secondary auditory cortex, we observed two clusters in the (B1) left- and (b2) right lateral occipital cortices (Post > Pre). (C) With the seed in the left frontal eye fields, one cluster was observed in the (C1) right temporal-occipital junction (Pre > Post). (D) With seed in the left primary motor cortex, we observed one cluster in the (D1) left insular cortex (Post > Pre). (E) With seed in the left primary visual cortex, we found one cluster in the (E1) left visual association cortex (Post > Pre). For details see Table 3. Color bars represent t-values. FWE = Family-Wise Error rate

### 3.5 Effect of the Multi-Modal Intervention in Glaucoma Patients

Next, we tested the changes in rs-FC in GL that underwent MMI using paired longitudinal t-tests. Significant changes were observed for eight seeds, i.e., the right dorsolateral prefrontal cortex, left midcingulate gyrus, left sensorimotor cortex, right primary motor cortex, right primary visual cortex, left secondary visual cortex, right primary auditory cortex, and left secondary auditory cortex (see Figure 6 and Table 3D). The right dorsolateral prefrontal cortex (BA9) showed significant changes in FC with the left superior frontal gyrus (p_FWE_ < 0.05, Post > Pre, cluster size = 34 voxels, p_FWE_ = 0.0074), left inferior frontal gyrus (p_FWE_ < 0.05, Post > Pre, cluster size = 29 voxels, p_FWE_ = 0.0202), and the right inferior temporal gyrus (p_FWE_ < 0.05, Post < Pre, cluster size = 26 voxels, p_FWE_ = 0.0379). The left MCG revealed significant changes in FC with the right precentral gyrus (p_FWE_ < 0.05, Post > Pre, cluster size = 31 voxels, p_FWE_ = 0.0119), and two clusters in the right cerebellum (p_FWE_ < 0.05, Post > Pre, cluster size = 32 and 29 voxels, p_FWE_ = 0.0097, 0.0180). We found significant changes in FC of left primary sensorimotor cortex (BA1) with the left inferior frontal gyrus (p_FWE_ < 0.05, Post > Pre, cluster size = 52 voxels, p_FWE_ = 0.0010). The right primary motor cortex (BA4) revealed changes in FC with the medial cuneal cortex (p_FWE_ < 0.05, Post > Pre, cluster size = 41 voxels, p_FWE_ = 0.0059). With the seed in the right primary visual cortex (BA17), we found significant changes in FC with the left inferior temporal gyrus (p_FWE_ < 0.05, Post > Pre, cluster size = 30 voxels, p_FWE_ = 0.0300). FC with the left secondary visual cortex (BA18) showed significant FC changes with the right postcentral gyrus (p_FWE_ < 0.05, Post Pre, cluster size =69 voxels, p_FWE_ = 0.000054). The right primary auditory cortex (BA41) showed increased FC with the left cuneal cortex (p_FWE_ < 0.05, Post > Pre, cluster size = 37 voxels, p_FWE_ = 0.0062) and the left secondary auditory cortex (BA42) showed increased FC with the left parieto-occipital cortex (p_FWE_ < 0.05, Post > Pre, cluster size = 41 voxels, p_FWE_ = 0.0048).

**Figure 6.**
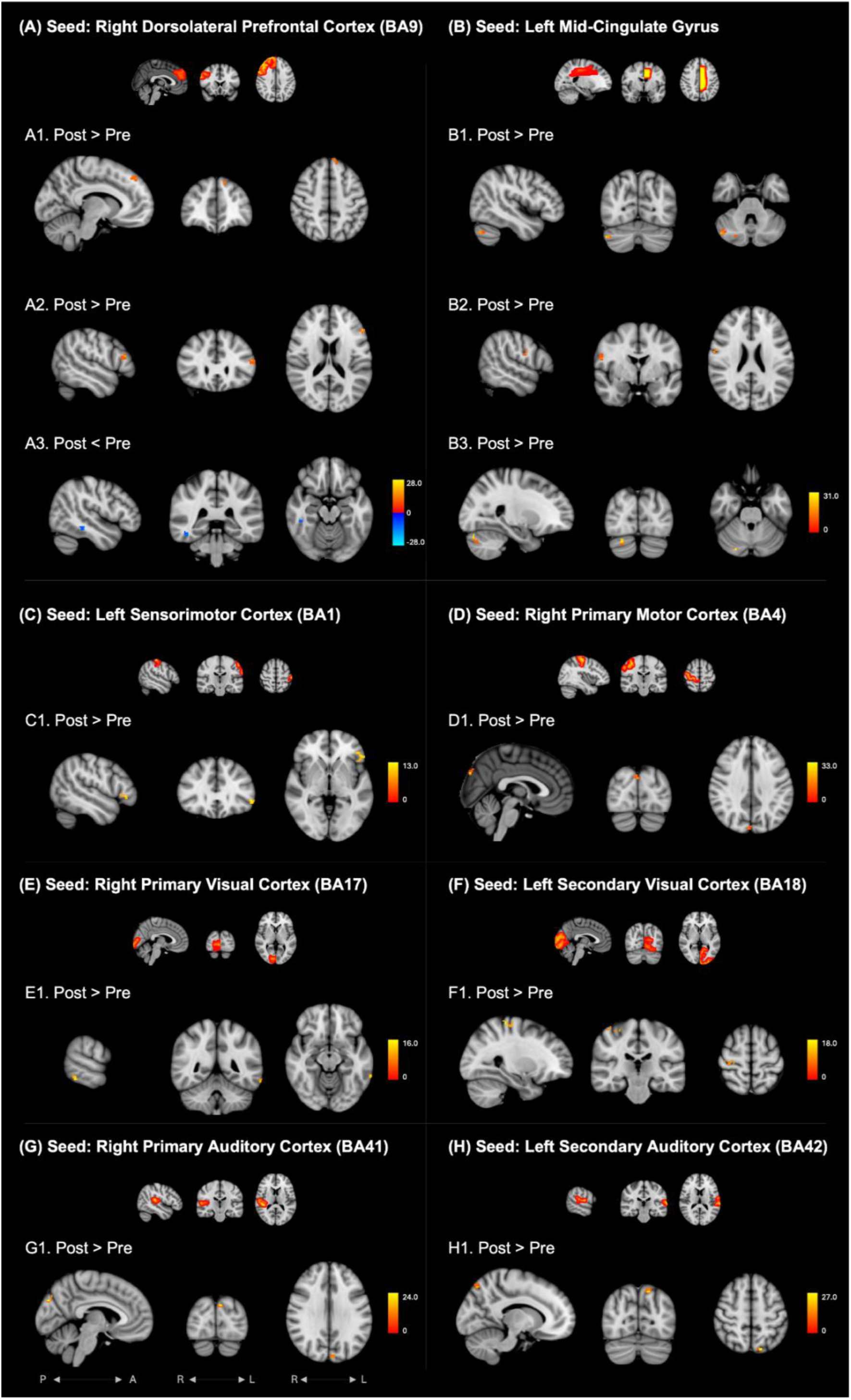
Significant differences in glaucoma patients before (Pre) and after (Post) the multi-modal intervention (pFWE < 0.05). (A) With seed in the right dorsolateral prefrontal cortex, three clusters were observed at (A1) left superior frontal gyrus (Post > Pre), (A2) left inferior frontal gyrus (Post > Pre), and (A3) right inferior temporal gyrus (Post < Pre). (B) With seed in the left midcingulate gyrus, three clusters were observed in the (B1) right cerebellum (Post > Pre), (B2) right precentral gyrus (Post > Pre), and (B3) right cerebellum (Post > Pre). (C) With seed in the left sensorimotor cortex, one cluster was found in the (C1) left inferior frontal gyrus (Post > Pre). (D) With seed in the right primary motor cortex, one cluster was observed in the (D1) medial cuneal cortex (Post > Pre). (E) With seed in the right primary visual cortex, one cluster was found in the (E1) left inferior temporal gyrus (Post > Pre). (F) With seed in the left secondary visual cortex, one cluster was observed in the (F1) right postcentral gyrus (Post > Pre). (G) With seed in the right primary auditory cortex, one cluster was found in the (G1) left cuneal cortex (Post > Pre). (H) With seed in the left secondary auditory cortex, one cluster was observed in the (H1) left parieto-occipital cortex (Post > Pre). For details see Table 3. Colorbars represent t-values. FWE = Family-Wise Error rate

## 4 Discussion

In this study, we investigated the effect of 12 weeks of MMI and UMI on FC in HC and GL in a two-arm randomized controlled design. Using rs-fMRI, we observed that participants who underwent MMI (HC or GL) showed significant changes in their FC with seeds in the visuo-motor pathway. Similar changes were not observed in the UMI counterpart. Moreover, there were differences in the effect of MMI between HC and GL.

### 4.1 Baseline Differences Before the Intervention

First, we tested for any baseline differences between the HC and GL cohorts before they performed the interventions. The comparison of FC in HC and GL has been studied in the past with heterogeneous outcomes and in different study populations (e.g. reviewed in Nuzzi et al., 2018, 2021). The present investigation is dominated by early disease stages and is therefore well comparable to that of Trivedi et al. (2019), who also focused on early glaucoma stages. Our comparison highlighted that FC in GL was higher between the auditory and visual association cortices compared to HC. This might be related to an increased reliance on multi-sensory integration as a compensation mechanism for loss of visual function in GL. In addition, we also observed increased FC between the auditory association cortex and the orbitofrontal cortex in GL compared to HC. The orbitofrontal cortex is generally involved in decision-making and reward processing (Wallis et al., 2007). Consequently, higher FC between the auditory and orbitofrontal cortices might be associated with increased involvement of auditory cues on routine decision-making activity in glaucoma patients. FC between the motor cortex and the FEF was also lower in GL compared to HC, which further suggests decreased dependence on visual processing in daily activities including motor control. Trivedi et al. (2019), also investigating people with early stages of glaucoma, reported FC between the visual cortex and the supramarginal areas in GL to be lower as compared to HC, an effect that appeared to be related to deficits in postural control. Consequently, our baseline comparisons indicate that our HC and GL cohorts were a good representative for the functional changes observed in early glaucoma.

### 4.2 Effects of the Unimodal Intervention

In the HC group that underwent UMI, an increase in FC of the cerebellum with MCG and FEF was found after training. The cerebellum is primarily involved in motor coordination, the MCG is implicated in decision-making and muscle control (Vogt et al., 2016), and the FEF facilitate oculomotor function. An increase in FC between these regions might indicate better motor coordination as a result of UMI in HC. Similar effects were also found by Macaulay et al. (2022) who observed an increase in the amplitude of low-frequency fluctuations (fALFF) of rs-fMRI in the cerebellum, middle temporal gyrus, and the inferior parietal cortex in older adults who underwent a 12-week intervention involving resistance training. Additionally, a longer duration of UMI could potentially have more widespread effects on brain function. For instance, participants in another study showed increased task-related activity in the middle temporal lobe, insula, and orbitofrontal cortex after 12 months of resistance training (Liu-Ambrose et al., 2012). Physical training has been shown to improve motor function and gait parameters in elderly participants (Fiatarone Singh et al., 2014, Puente-Gonzalez et al., 2021), also in cases of mild cognitive impairment (Dove et al., 2020) and Parkinson’s disorder (Allen et al, 2011). Surprisingly, we did not find significant changes in the GL who underwent UMI. The lack of significant longitudinal effects of UMI in GL patients suggests that they might either be less pronounced in this cohort or require larger sample sizes to reach significance.

### 4.3 Effects of the Multimodal Intervention

In contrast to UMI, participants that underwent the MMI performed multi-sensory integration tasks (as a part of ‘Life Kinetik’) in addition to basic resistance training. To date, the effects of Life Kinetik on resting state FC have been reported in a single study, employing Life Kinetik exclusively in a healthy cohort comparing younger participants (mean age 48 years) than the present study population (Demirakca et al., 2016). In their study, it was found that FC between the auditory and somatosensory cortices was increased after training. They also reported increased FC between the visual and motor regions. FC of the ventral anterior cingulate cortex with the prefrontal cortex, visual areas, and FEF was also affected. Another study that employed a similar multi-modal intervention scheme (involving cognitive tasks, counseling, and Tai Chi) reported an increase in resting-state FC between the prefrontal cortex and medial temporal lobe after training that correlated with improvement in cognitive measures (Li et al., 2008). In our study, we employed “Life Kinetik” combined with resistance training, and we investigated two cohorts of participants (HC and GL, both ≥ 60 years of age) that underwent the intervention. In both HC and GL, we observed considerably more FC changes after MMI compared to UMI. In our cohort, we found similar effects in the FC between the visual and auditory cortices after MMI in HC. However, in contrast to Demirakca et al. (2016), we did not find a widespread effect of MMI on FC with the somatosensory and motor cortices.

Among the array of longitudinal effects of MMI, certain patterns showed similarities across the HC and GL cohorts. In both HC and GL, we found that the FC between the primary visual cortex and the higher visual areas was increased after MMI. In HC, this increase was observed in FC between the primary and secondary visual cortex (V2). Similarly, in GL, FC between the primary visual cortex and the posterior parts of the inferior temporal gyrus was increased after MMI. This indicates that MMI increased the FC in both groups along the visual pathway, specifically along the ventral stream. Improved FC along the ventral visual stream could be indicative of improved ability of object recognition or a better ability to associate sounds/words with visual input (O’Neil et al. 2014, Li et al. 2018).

We also found that the FC between the auditory and visual areas was increased in both HC and GL after MMI. In HC, we found that FC between the BA41 and the parieto-occipital cortex showed an increment, whereas in GL, the superior parts of the cuneal cortex showed increased FC with BA41 after MMI. The visual and auditory association areas also showed increased FC in both groups after MMI. In HC, FC between the auditory association cortex (BA42) and the right and left lateral occipital cortices showed an increment after MMI. Similarly, in GL, we observed that the FC between the auditory association cortex and the parieto-occipital cortex increased after MMI. Increased FC between regions of the auditory and visual processing pathways suggests improved integration of sensory information from both modalities. The similarities indicate that the MMI had a consistent impact on the audio-visual integration in the participants, both HC and GL. These changes could be linked to the multi-sensory nature of the tasks involved in MMI that required simultaneous processing of auditory cues, visual object tracking, and motor planning. A similar increase in FC between secondary auditory and visual cortices was also observed by Demirakca et al. (2016), although the effect of training on audio-visual FC was not as pronounced in their cohort. This could be explained by the higher dosage of training in our study (total 24 hours over 12 weeks) compared the study of Demirakca et al. (2016) (total 11 hours over 13 weeks). A higher exercise volume in our study could have helped accentuate the task-relevant changes and attenuate the changes related to generally increased motor activity due to the intervention.

In addition to the consistent impact of MMI on audiovisual integration in both HC and GL, we also observed some changes that were unique to HC or GL. In HC, we found that the FC of the auditory cortex with the parieto-occipital areas and the hippocampus was increased after MMI, suggesting greater involvement of auditory information in spatial awareness and memory formation. Demirakca et al. (2016) also reported a similar increase in FC between auditory and parietal regions. We also found an increase in FC between the motor cortex and the insular cortex after MMI, accompanied by reduced FC between the FEF and the temporal-occipital junction. The latter finding contrasts with the reports by Demirakca et al. (2016), as they found increased coactivation of the FEF with the visual cortex after MMI in HC. It is difficult to compare the nuances of FC changes as they could be highly dependent on the differences in the study population and other design-specific differences.

In contrast to HC, FC in GL was altered after MMI in multiple regions in the frontal cortex. We observed increased FC between the inferior frontal gyrus, dorsolateral prefrontal cortex, superior frontal gyrus, and the inferior temporal gyrus. These frontal regions are primarily involved in attention, multi-tasking, decision-making, as well as error and reward processing (Rushworth et al. 2011, Markett et al, 2022). An increase in FC of frontal regions with the regions in the ventral visual pathway indicates greater involvement of the attention and decision-making circuits in object recognition and detection in GL patients after MMI. Deficits in FC between the visual cortex and the inferior/superior frontal gyri have been reported in GL in multiple studies (Nuzzi et al., 2018, 2021). Based on our findings, MMI seems to target this deficit in GL patients. Assessment of visual functions related to the ventral pathway (object recognition, detection, and tracking) could shed more light on these FC changes in GL. We also observed that, like UMI in HC, the FC between the cerebellum, precentral gyrus, and the MCG was higher after the MMI in GL. This might be associated with the resistance-training component of MMI. Increased FC between the cerebellar and motor cortices was also reported by Demirakca et al. (2016) in their cohort of HC after Life Kinetik. Increased FC in circuits involved in motor coordination, coupled with strengthened FC between the visual and frontal cortices could indicate improved sensori-motor coordination in GL who underwent MMI. Future studies with a larger sample size would enable relating fMRI-based findings to scores in visually-guided motor coordination tasks, helping associate the observed FC changes with practical outcomes.

### 4.4 Comparing UMI and MMI

It has been observed in elderly HC that a combination of motor and cognitive training could be more effective in enhancing cognitive and physical function (Dove et al, 2020; Schoene et al, 2014). To our knowledge, the present study is the first study investigating FC changes after MMI and UMI. We expected to find rs-FC changes corresponding to aforementioned behavioral outcomes in our cohort. It is evident from our findings that MMI targets a larger set of functional networks in both HC and GL than UMI. In participants who underwent UMI, we observed the effect of training primarily in the regions involved in motor planning and execution. On the other hand, the impact of MMI was observed in audio-visual networks, frontal attention networks, parietal regions, as well as the networks involved in motor planning and execution. These distinctions might be associated with the differences in the nature of tasks involved in MMI and UMI. Our findings extent those reported by Demirakca et al. *(2016)*, highlighting the widespread changes after MMI that are not observed after UMI. However, a future follow-up study with a larger sample size would be required for systematic statistical comparison of the intervention and group factors, along with their interaction effects. Moreover, longitudinal quantitative assessment of task performance measures would be essential in testing the real-life impact of the intervention.

In multiple studies focusing on the elderly, it has been reported that motor-cognitive training can increase motor performance, gait measures, and cognitive function (Wollensen et al., 2014). A recent study comparing the effects of resistance training and combined resistance-cognitive training found that both interventions improved walking abilities and gait parameters, but larger improvements in cognitive and motor dual-task abilities were found after the combined training (Wang et al., 2024). Moreover, a multimodal visuo-motor integration training was found to be effective in improving memory, visual-motor integration, and memory self-efficacy in elderlies (Kim et al., 2017). Systematic reviews with meta-analyses by Wollensen et al. (2020) and Schoene et al. (2014) concluded that motor-cognitive dual-task interventions should improve global cognitive function and inhibitory control in elderlies. Multimodal training has also been shown to improve mobility and executive functions in the elderly with mild cognitive impairment (de Oliveira Silva et al, 2019). Our findings add to this evidence that MMI has a broader impact on the FC in elderly HC as compared to UMI. Further, given the visuo-motor dysfunction in glaucoma, we suggest that MMI could be an effective strategy to increase FC in GL patients, which positively influence visuo-motor function.

### 4.5 Limitations and Future Scope

This study aimed to understand the effects of MMI compared to UMI in HC and GL. The sample size of the cohorts was, due to pandemic limitations (COVID-19), smaller than initially intended (Xue et al. 2020). As a consequence, unpaired statistical tests for the subgroups were deemed unreliable. Future investigations with a much larger sample size would enable more complete and reliable statistical testing using a 2^3^ factorial model, such that the interaction of group and intervention type could also be studied more systematically. Such an approach should also test the relation of the resting state FC effects to task-based fMRI paradigms and behavioral, cognitive, and motor-coordination tests, e.g., by conducting quantitative correlation analyses between FC changes and task performance scores. Such analyses could inform us about the behavioral relevance of FC outcomes of MMI and UMI in HC and GL.

## 5 Conclusion

We conducted a two-arm randomized controlled study to investigate the impact of MMI and UMI on rs-FC in HC and GL. After UMI, we observed a longitudinal increase in FC in the networks involved in motor planning and coordination only in HC. Whereas, after MMI, we observed widespread effects in both HC and GL. In both HC and GL, MMI impacted FC in the visual pathway, audiovisual networks, and visuo-motor networks. In GL, MMI also increased FC in the frontal attention and decision-making networks. Overall, the findings suggest a widespread effect of MMI in both HC and GL on rs-FC, motivating future studies with larger sample sizes and task performance assessments to comment on the real-life impact of the intervention.

## 7 Data Availability Statement

Data shall be made available upon reasonable request. Data processing was done using standard pipelines in SPM12 and CONN toolboxes without custom scripts.

## 8 Acknowledgements

We thank the study participants for their support of the study.

